# Validating sporicidal efficacy of ultrasound probe high-level disinfection devices in clinical settings

**DOI:** 10.1101/2025.02.03.25321618

**Authors:** David Bellamy, Karen Vickery

## Abstract

**Introduction:** In accordance with AS5369:2023 and ISO 15883-1:2024 Standards, in-field validation testing of automated high-level disinfection (HLD) devices in clinical settings is necessary to qualify their performance. Responding to reports that some ultraviolet-C (UV-C) devices were failing to achieve sporicidal efficacy during routine in-field validation, we evaluated the sporicidal efficacy of these devices via a performance qualification test.

**Methods:** Sporicidal efficacy was assessed using commercially available stainless steel biological indicators (BIs) inoculated with 10^6^ *Geobacillus stearothermophilus* spores (ATCC^®^ 7953). BIs were clamped in top and bottom locations inside chambers of devices [UV-C light-emitting diode (LED), UV-C lamp and hydrogen peroxide (H_2_O_2_) mist]. BI test conditions included packaged, unwrapped and non-flamed and unwrapped and flame sterilised on the clamped coupon end. Results were evaluated on a pass (no growth) or fail (growth) basis.

**Results:** The results showed that the UV-C LED device failed to inactivate spores in all tested positions and conditions (n=18). The UV-C lamp device passed 2/6 tests in the flamed condition but failed all other tests (n=12). The H_2_O_2_ mist system passed all tests, inactivating spores for all conditions and chamber positions (n=18).

**Conclusion:** Our findings show that despite claiming sporicidal efficacy, the UV-C devices both failed in-field validation tests using bacterial endospores. These results indicate that the UV-C devices were not sporicidal in these tests. The H_2_O_2_ mist device was the only system in this study that passed all tests, achieving sporicidal efficacy.

## Introduction

The appropriate reprocessing of reusable medical devices before individual patient use is essential for the prevention of infection transmission (1). Ultrasound probes that contact non-intact skin or mucous membranes during patient procedures are classified as semi-critical devices and, at a minimum, require high-level disinfection (HLD) (2,3).

There are three automated systems intended to provide HLD of ultrasound probes currently in clinical use in Australia and New Zealand (ANZ). All of these devices claim efficacy against bacterial endospores according to their product literature (4-6). The Therapeutic Goods Administration (TGA) standards for sporicidal efficacy require demonstrated inactivation of 10^6^ spores (7) and sporicidal products should be able to achieve a 6-log reduction of any bacterial endospore, including species such as *Geobacillus stearothermophilus*. The three devices available in ANZ utilise ultraviolet-C (UV-C) light-emitting diodes (LEDs), UV-C lamps, or hydrogen peroxide (H_2_O_2_) mist in enclosed systems.

Routine in-field validation testing of disinfection devices in clinical use is required under both Australian (AS 5369:2023) (8) and International Standards (ISO 15883-1:2024) (9). Performance qualification reflects one stage of in-field validation and requires testing the biological inactivation of microorganisms using a biological indicator (BI) test (AS 5369:2023) (8).

Performance qualification of automated disinfection devices is required after installation, upon servicing and requalification annually under Australian standards (AS 5369:2023) (8). We became aware of data showing that some UV-C devices were failing to achieve sporicidal efficacy during routine validation in hospitals located in Australia and set out to test the efficacy of these devices ourselves in a rigorous in-field validation process using BIs.

We assessed the sporicidal efficacy of the disinfection processes through the destruction of 10^6^ bacterial endospores (*G. stearothermophilus*), consistent with TGA requirements (7). These BIs were chosen on the basis that manufacturers of all three technologies evaluated in this study claim sporicidal efficacy (4-6).

## Materials and methods

### Automated disinfection devices

The performance of three commercially available automated disinfection devices was evaluated in clinical settings across ANZ. The devices included two UV-C systems [UV-C LED (Lumicare ONE, Lumicare) and UV-C lamp (Hypernova Chronos^®^, Germitec)] of different wavelengths and dimensions, and a hydrogen peroxide mist system (H_2_O_2_ mist: trophon^®^2, Nanosonics) (Table 1). All devices were operated according to the manufacturer’s Instructions for Use (IFU).

**Table 1.**
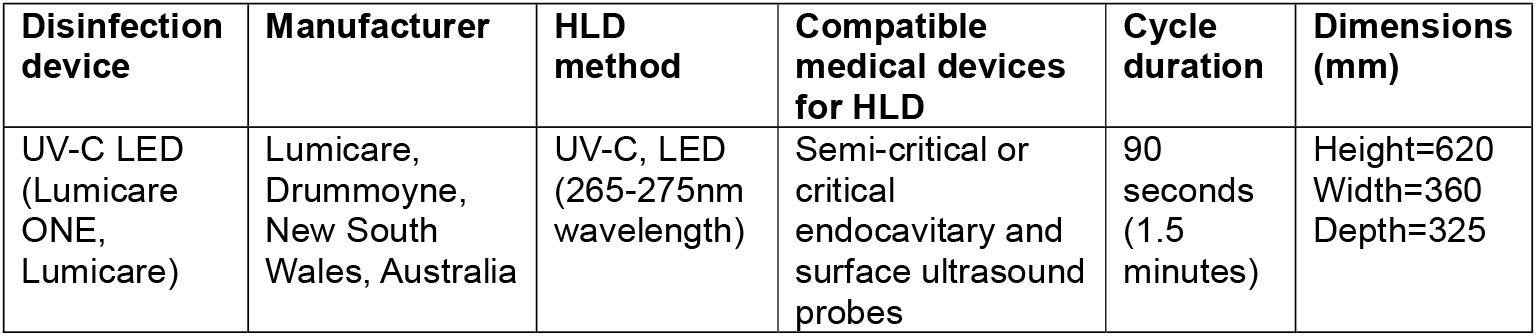
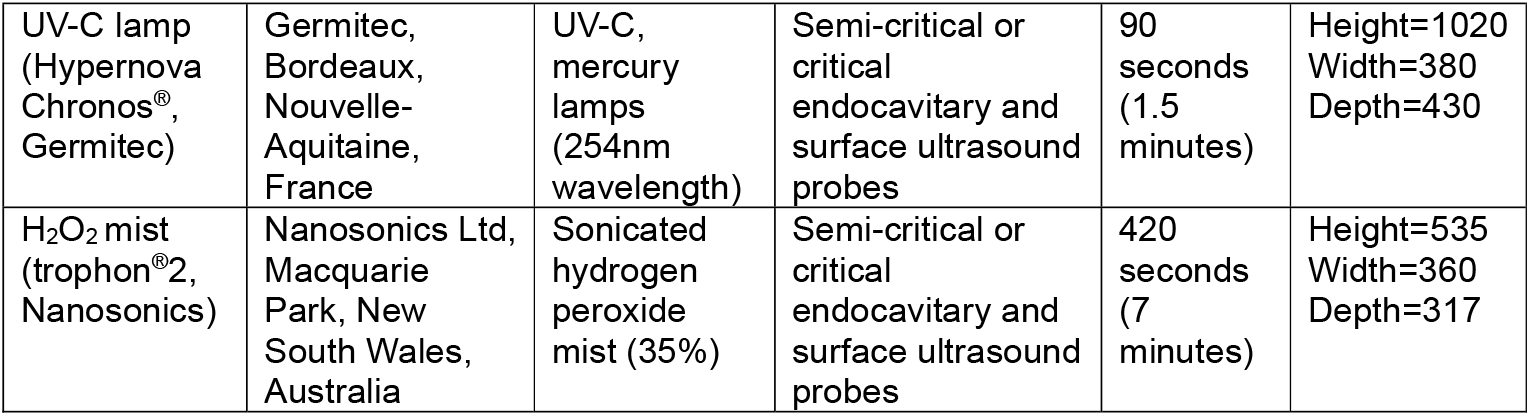
Specifications of the assessed disinfection devices.

### Procedures

BIs containing 10^6^ *G. stearothermophilus* (ATCC^®^ 7953) spores inoculated onto stainless steel coupons (dimensions: 34.0mm × 7.0mm × 0.8mm) (Terragene^®^, BT93/6) enclosed in a glassine package/ Tyvek^®^ (dimensions: 25mm × 70mm) were used.

Given that UV-C light has limited penetration and is subject to shadowing, we opted to test three conditions; 1) The BI in its typical glassine packaging/ Tyvek^®^; 2) The BI with packaging removed and; 3) The BI with packaging removed plus flame sterilisation of the coupon end which was later clamped (Figure 1A-1C).

**Figure 1.**
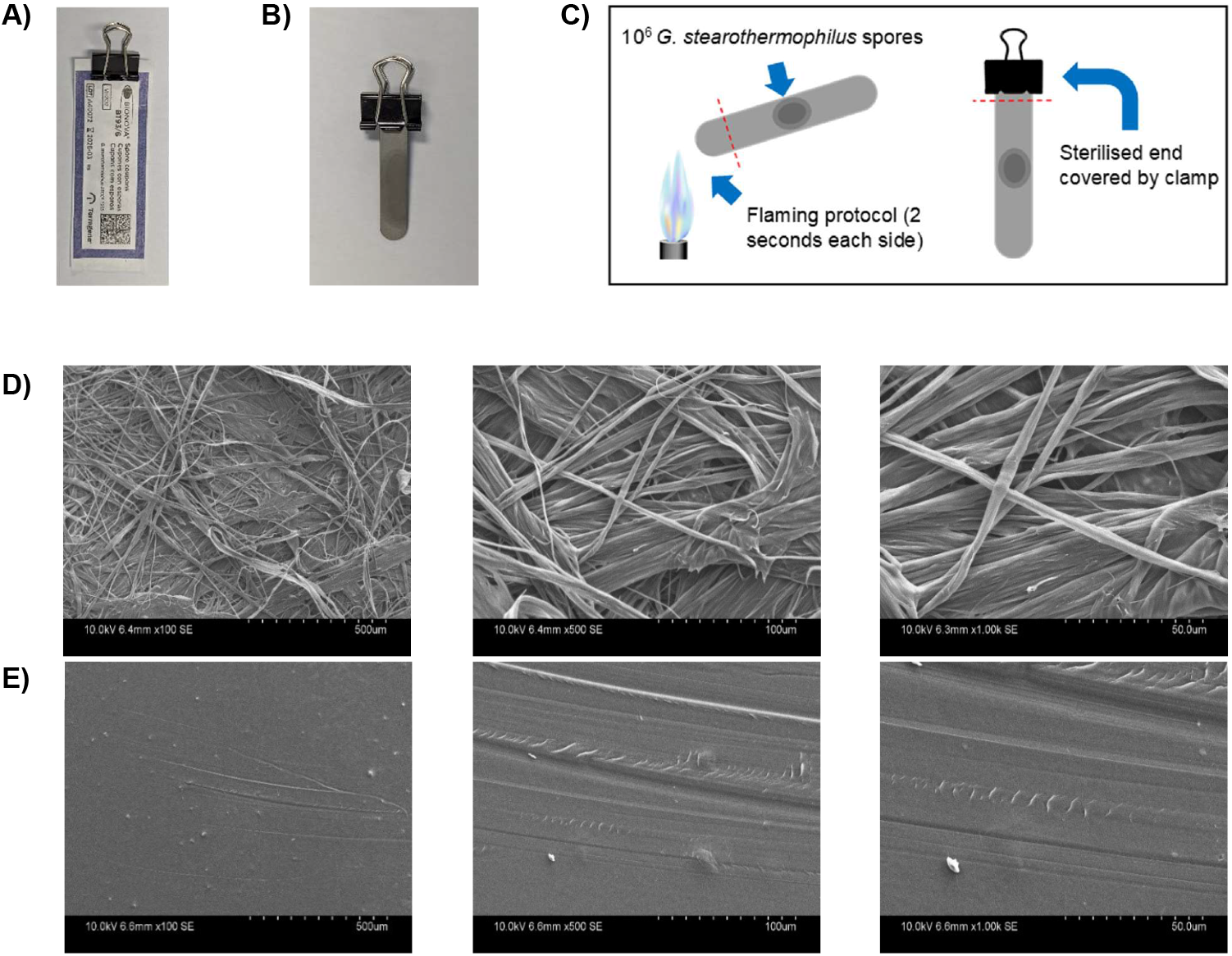
Three BI test conditions and scanning electron microscopy (SEM) images of BI (Terragene^®^, BT93/6) glassine packaging/ Tyvek^®^. **A)** BI coupon contained within a glassine package/ Tyvek^®^. **B)** BI coupon without packaging. **C)** A diagram of the BI with a flamed sterilised coupon end. **D)** SEM images showing the structure of the high-density polyethylene side of the BI package is fibrous in nature, allowing the penetrability of very small particles. **E)** SEM images showing that the structure of the plastic side of the BI package is non-permeable in nature with no visible pores.

BIs corresponding to the test conditions were suspended in the disinfection chambers via a sterilised clamp which was attached to one end of the coupon or packaging. Due to potential shadowing effects on the coupon from the clamp, a flame sterilisation technique of the clamped coupon end was employed to account for any shadowing, where the shadowed areas would not be accessible to UV-C radiation. This was performed by flame sterilising 5mm (0.5cm) of the BI coupon ends for 2 seconds on each side of the same end (1 minute cooling time in between) (see S1). The sterilised end of the coupon was then clamped onto the coupon holder inside the respective disinfection device.

For the non-flamed BI test condition without glassine packages/ Tyvek^®^ (stainless steel BI coupon only), BIs were aseptically removed from the packaging and directly clamped onto the holder. For the BIs tested with glassine packages/ Tyvek^®^, the edge of the package was clamped and suspended inside the disinfection device using the holder (BI coupon itself was not clamped when inside the package).

BIs were positioned, with the inoculation site facing outwards, in the top and bottom positions inside the disinfection chamber to represent locations within the chamber corresponding to where a clinical probe would be placed (Figure 2). Three replicates were used for each test condition for both top and bottom positions in the chamber, totalling 6 replicates per device. The holder was suspended in each disinfection device and exited from the chamber via the cavity located at the top where an ultrasound probe cable would typically emerge.

**Figure 2.**
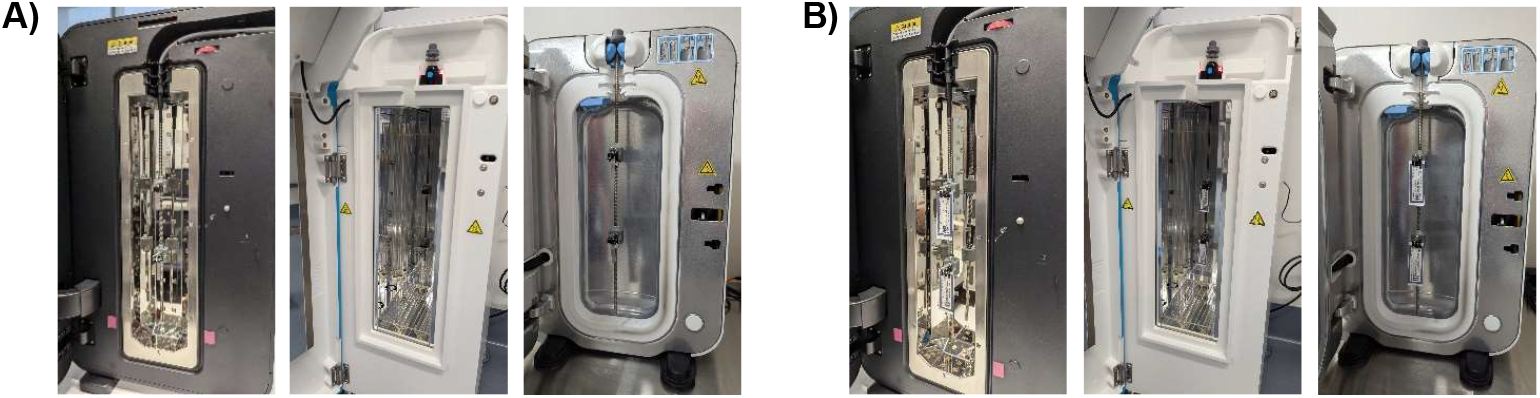
Set up of the BIs clamped onto the holder positioned in the top and bottom of the tested disinfection chambers. **A)** BI coupons without glassine packages/ Tyvek^®^ (flame sterilised coupon end or non-flamed). **B)** BI coupons contained within glassine packages/ Tyvek^®^.

A standard disinfection cycle was carried out according to the disinfection device manufacturer’s IFU. Three cycles were conducted per test condition, with each BI placed in the top and bottom positions (2 BIs) for each cycle, for a total of 6 replicates per condition. After the disinfection cycle, each BI coupon was aseptically collected into suitable culture media according to the manufacturer’s IFU.

Three positive controls were included for both the flamed and non-flamed test conditions (total of 6 positive controls per device) and were prepared by aseptically transferring the BI directly into the corresponding culture media vial. The inclusion of positive controls ensured that the incubation process did not contribute to a false-negative result.

All vials were then shipped to an accredited, independent test laboratory and incubated and assessed for growth/no growth according to the manufacturer’s IFU. Growth indicated failure of the test, and no growth indicated a pass result.

Data analysis was performed using R-Studio software (version 4.1.1). To examine the relationship between disinfection devices and test conditions (flame sterilised, non-flamed, glassine package/ Tyvek^®^), a Fisher’s exact test was performed to assess independence. A p-value less than 0.05 indicated statistical significance.

## Results

The study assessed a total of 72 BIs across the three disinfection devices (UV-C LED, UV-C lamp and H_2_O_2_ mist), three test conditions (flamed, non-flamed and glassine package/ Tyvek^®^), with 18 positive controls for the flamed and non-flamed conditions. The H_2_O_2_ mist device exhibited the highest sporicidal efficacy with a 100% pass rate across all tested conditions and chamber positions overall (18/18 replicates) (Table 2).

**Table 2.**
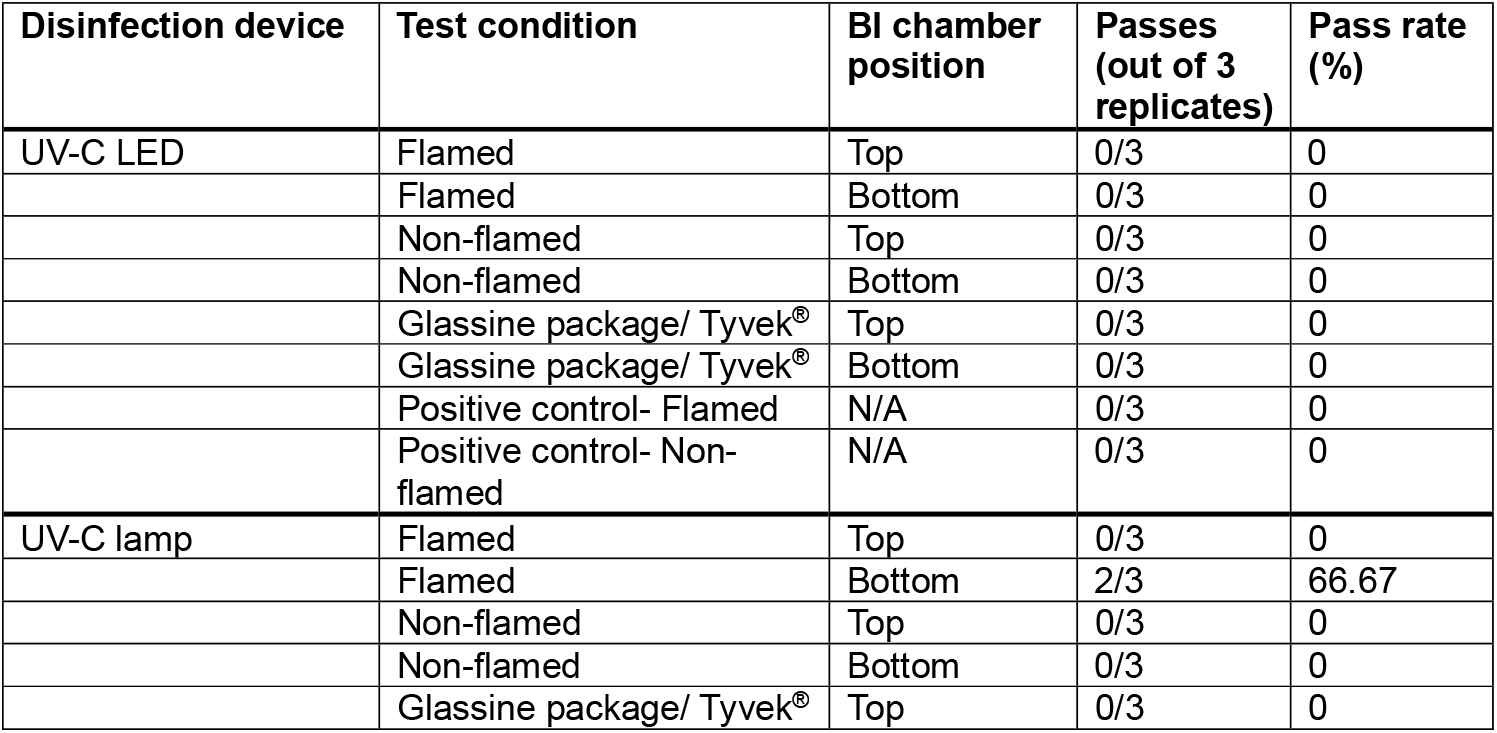
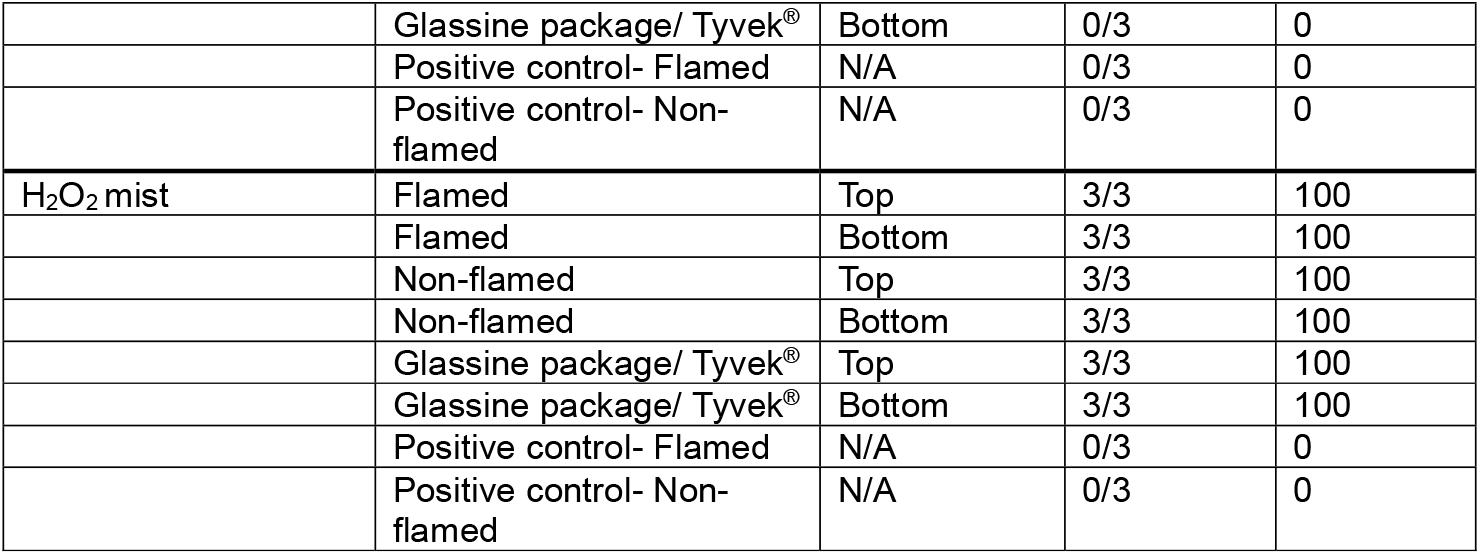
BI testing results for each of the evaluated disinfection devices.

The UV-C lamp demonstrated partial efficacy under the flamed condition, with a 66.67% pass rate (2/3 replicates) in the bottom position, but a 0% pass rate (0/3 replicates) in the top position, with a 33.33% pass rate for the overall flamed condition (2/6 replicates). The UV-C lamp failed to inactivate spores across any of the non-flamed and glassine/Tyvek^®^ package conditions (0/12 replicates), with an 11.11% overall pass rate across all tested conditions (2/18 replicates).

In contrast, the UV-C LED device failed in inactivating spores across all tested conditions and positions (0/18 replicates) (Table 2).

Statistical analysis under all test conditions revealed significant differences in the pass/fail rate between the devices, χ^2^(2, N=6), p<0.0021, p<0.0002, p<0.0002, for flamed, non-flamed and glassine package/ Tyvek^®^ conditions, respectively. All positive controls demonstrated spore growth, validating incubator function and spore viability.

## Discussion

The in-field validation results showed that the H_2_O_2_ mist system was effective at sporicidal disinfection in all cases, while the UV-C LED and UV-C lamp systems were largely ineffective. Two of the six replicates were successfully disinfected by the UV-C lamp system when using a BI without packaging with the flame sterilised end. However, these successful tests only occurred in the bottom chamber position, while all replicates in the top position failed.

The UV-C lamp device performed better than the UV-C LED device, achieving sporicidal efficacy for 2 of 6 flame sterilised BIs, while the UV-C LED device was ineffective in all tested cases, irrespective of the BI condition or chamber position (Table 2). On reviewing the literature, we identified a manufacturer-funded study on the UV-C LED system. This study reported a >7-log_10_ reduction of *Bacillus subtilis* spores after undergoing UV-C disinfection (10). In the previous study, the spores were spread over a very large area compared to the standard BIs used in the present study. The lower density of the inoculum could pose a lesser challenge and may explain the differences in results to the present study.

The differences observed between the UV-C devices may be attributable to the varied wavelength, dose, temperature, reflectivity of the chamber wall material and configuration of light sources in the chamber (11,12,13). Overall, the results show that the automated UV-C disinfection devices did not consistently achieve sporicidal inactivation. As the validation of disinfection devices is a requirement according to Australian and international standards (8,9), automated disinfection devices, particularly UV-C disinfection devices, should be successfully validated to ensure they are meeting claims such as sporicidal efficacy.

In order for a UV-C device to achieve a pass result under these test conditions, it must deliver a sufficient dose of UV-C radiation to the BI to effectively inactivate all spores present. It is evident in the study that the UV-C devices were unable to deliver an adequate dose to the relatively uniform, flat surface of the BI coupons to achieve sporicidal efficacy. While there are several known factors that can impact sporicidal efficacy of UV-C based technologies, the dose received by an area of a probe is dependent on the irradiance level on the target surface and the duration of exposure. As the duration of exposure is dictated by the pre-set cycle time, it is critical that the irradiance level on the surface is sufficient to inactivate the spores.

UV-C requires a direct path of light to disinfect the target surface, with intensity falling with the square of distance from the target (inverse square law), impacting the amount of UV-C radiation that is received (11). As such, the distance of the BI from the light source may have resulted in a sub-efficacious dose. Mercury lamps have been shown to exhibit less variability in irradiance distributions compared to UV-C LEDs at shorter distances from the surface plane (14). Given that areas closer to the source of light receive greater irradiance (11), it may be the case that the configuration and distance of light sources in the bottom location of the UV-C lamp chamber were better able to irradiate an appropriate dose onto the BIs, although inconsistently. This finding, coupled with the failing of all the BIs located in the top position, demonstrates that irradiance is not equally distributed within the entire chamber.

In addition to the above considerations, the dosage of UV-C irradiated onto the coupon surface may have been affected by variables such as the surface topography and potential micro-shadowing on the surface of the coupon (15). The potential for shadowing effects may impact disinfection efficacy, which increases the risk of microorganisms surviving in areas of low irradiance (11). As the potential for large variation in irradiance distribution has been noted, testing is needed to demonstrate efficacy in areas of least irradiance (16). Despite employing flame sterilisation on the clamped coupon end in the present study to account for potential shadowing in the clamped area, UV-C was still unable to effectively inactivate the spores, even in non-shadowed areas on the coupon. These limitations of uneven irradiance distributions and potential shadowing underscore the challenges of UV-C disinfection devices in achieving adequate disinfection, particularly for complex surfaces.

Several recent studies investigating the use of UV-C for the disinfection of semi-critical devices found that the HLD criteria were not consistently met as microorganisms remained on the devices following HLD (10,17-20), suggesting that UV-C technology may not be able to deliver effective HLD in all cases.

The current study found that the H_2_O_2_ mist system passed all tests, inactivating spores across all replicates, in all conditions tested. This demonstrates that the H_2_O_2_ mist device met the TGA standards for sporicidal efficacy by achieving the inactivation of 10^6^ bacterial endospores (7). These findings may be attributable to the ability of the H_2_O_2_ mist device to distribute H_2_O_2_ mist across the carrier surface which ensures greater surface coverage and spore inactivation, even in shadowed areas around the clamp. These data align with existing literature on the efficacy of the H_2_O_2_ mist system for sporicidal disinfection (1).

Interestingly, the H_2_O_2_ mist device inactivated the spores on the BIs contained in glassine packages/ Tyvek^®^, whereas the UV-C devices were ineffective in penetrating the packages to inactivate the spores. This demonstrates that the H_2_O_2_ mist device provided more effective penetration of difficult-to-reach areas, such that there is adequate saturation and coverage of the entire surface of a contaminated device, and thus sporicidal efficacy. While the transparent plastic side of the glassine package/ Tyvek^®^ is non-permeable to molecules, the glassine package/ Tyvek^®^ side of the package contains high-density polyethylene fibres designed to allow penetration of gases and very small particles (Figure 1D and 1E), while maintaining an effective barrier to microorganisms such as bacterial cells (≈µm diameter) and fungal spores (>1µm diameter) (21). Given that the glassine package/ Tyvek^®^ is hydrophobic in nature, it is apparent that the H_2_O_2_ mist effectively permeates the glassine package/ Tyvek^®^ to inactivate the spores on the BI coupon.

A limitation of the study is that BI coupons containing different surface materials and bacterial spores were not investigated. However, a standardised and commercially available BI test was employed in the study and these BIs are used widely to validate the sporicidal efficacy of disinfectors or sterilisers. Furthermore, the implication and claims of sporicidal efficacy made by the manufacturers represent that the devices used within this study would demonstrate efficacy regardless of the BI chosen. In this regard, it is worth noting that the spores used within this study are not worst-case spores for UV-C technology, and therefore additional testing with worst-case microorganisms is warranted. In addition to the above, there may have been some variation in the positioning of the BI coupons in the top and bottom locations of the disinfection chamber across the devices due to differences in device sizes. Future studies on automated HLD devices in clinical use, including quantification methods such as colony forming units with testing directly on ultrasound probe surfaces, may be beneficial to corroborate the current findings on sporicidal efficacy.

In conclusion, the results show that the H_2_O_2_ mist device passed the sporicidal tests, whereas the UV-C devices failed most or all of the tests, depending on the system tested. Failure of in-field validation in these cases corresponded to a failure in sporicidal efficacy. This raises questions as to whether UV-C devices are able to consistently ensure effective inactivation of spores on ultrasound probes in clinical settings. Periodic and successful in-field validation of automated disinfection devices is crucial to protect the health and safety of healthcare professionals and patients.

## Supporting information

Flame Method

## Data Availability

All data produced in the present work are contained in the manuscript

## Ethical considerations

Ethical approval and informed consent were not required as there were no patients or volunteers involved in this study.

## Authorship statement

D.B: conceptualisation, writing – review and editing. K.V: conceptualisation, methodology, writing – review and editing.

## Acknowledgements

The authors would like to thank the hospitals for use of their facilities.

## Supplementary information

S1. Flame sterilisation method validation.

