## Supplementary material for "Validating sporicidal efficacy of ultrasound probe high-level disinfection devices in clinical settings": Flame Method

### S1. Flame sterilisation method validation

#### A. Procedure for wrapping of flamed BI (Terragene®, BT93/6)

1. Dry a Tryptic soy agar (TSA) plate at 55˚C for at least 1 hour, with the lid a little off to remove condensation.
2. Aseptically open the packaging of BI stainless steel coupon (Terragene®, BT93/6)
3. Use sterilised forceps to hold the BI coupon in a vertical position (flat surface of the coupon is vertical with respect to the lab bench).
4. Flame sterilise 0.5cm of the top end of the BI coupon (in vertical position) in the blue flame of the Bunsen burner for 2 seconds.
5. Allow coupon to cool to room temperature for 1 minute.
6. Turn the BI 180˚ and flame sterilise the opposite side of the BI in the blue flame of the Bunsen burner for 2 seconds.
7. Allow coupon to cool to room temperature for 30 seconds.
8. Wrap the non-flamed part of the BI with sterile parafilm.
9. Place the BI coupon onto the dried TSA plate (spore spot side down).
10. Incubate TSA plate at 55˚C, with the lid a little off, to avoid building up of the condensation, put in a box with closed lid.
11. View the plates at 24 hours.

**A)**

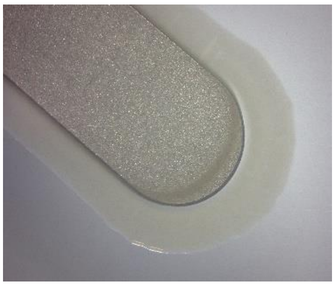

**B)**

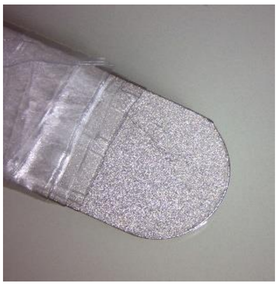

**Figure S1.1. A)** The control BI with a flamed top end without parafilm wrapping on the non-flamed area showed growth of *G. stearothermophilus* spores around all areas of the coupon. **B)** The BI flamed at the top end with the non-flamed area wrapped in parafilm showed no growth of *G. stearothermophilus* spores.

#### Procedure for wrapping of non-flamed BI (Terragene®, BT93/6)

1. Dry a TSA plate at 55˚C for at least 1 hour, with the lid little off to remove condensation.
2. Aseptically open the packaging of stainless steel coupon (Terragene®, BT93/6).
3. Use sterilised forceps to hold the BI coupon.
4. Wrap the same part of the BI as done in step A8 with sterile parafilm.
5. Repeat steps A9 to A11.

**A)**

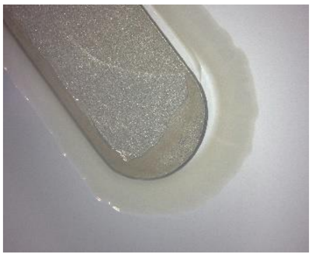

**B)**

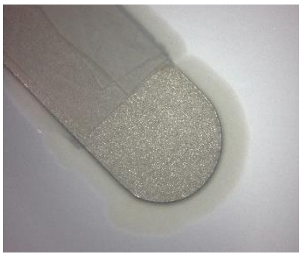

**Figure S1.2. A)** The control non-flamed BI without wrapping showed growth of *G. stearothermophilus* spores around all areas of the coupon. **B)** The non-flamed BI with the same area as Figure S1.1B wrapped in parafilm showed growth of *G. stearothermophilus* spores around the wrapped and non-wrapped areas.
